# Risk beliefs, intensive digital information and demand for a new preventative health product in public clinics: Evidence from an experiment in Zimbabwe

**DOI:** 10.64898/2026.06.15.26355642

**Authors:** Ranjeeta Thomas, Matteo M. Galizzi, Louisa Moorhouse, Phyllis Mandizvidza, Freedom Dzamatira, Simon Gregson

**Affiliations:** Department of Health Policy, London School of Economics and Political Science, London, United Kingdom; Department of Psychology and Behavioural Science, London School of Economics and Political Science, London, United Kingdom; MRC Centre for Global Infectious Disease Analysis, School of Public Health, Imperial College London, United Kingdom; Biomedical Research and Training Institute, Harare, Zimbabwe

**Keywords:** risk beliefs, health product, preventative health care, risky behaviour

## Abstract

Demand for preventative health care is weak in low-income settings. In a field experiment in a low-income, high-risk setting, we evaluated whether demand for a new bio-medical preventative health product, offered free at public health clinics, responds to digital feedback-based intensive information on health risks and benefits of prevention along with a clinic referral enabling access to the product. In our sample of women aged 18-24 years, we find a large correction in risk beliefs sustained six months after the intervention. Against a background of very low baseline usage, within six months we find a 5.8 percentage point increase in take up of the prevention method, a level of uptake which is very large relative to the control group. Reassuringly, there is no meaningful difference in up-take amongst baseline high- risk and low-risk individuals.

## 1. Introduction

Demand for preventative health care is generally poor in low-income countries. Even when free, demand may be low due to – inaccurate prior beliefs around health risks and benefits of new preventative methods; high immediate non-monetary costs and limited agency to mitigate risks; and the extent to which an individual trades off future benefits against immediate costs (present bias) (Dupas & Miguel, 2017; Kremer et al., 2019). In this study, we evaluate whether the demand for a *new*, publicly provided preventative bio-medical health product responds to an intervention that combines digital feedback-based information to correct inaccurate risk beliefs with a clinic referral for the product. The preventative health product of interest is Pre-exposure Prophylaxis for HIV (PrEP) which at the time of our study was not yet available at local public clinics but was being planned for a gradual national roll out as a new preventative health product for at-risk individuals.

We study demand responses of young adult women (18-24 years) in Zimbabwe, an age group at high risk of acquiring sexually transmitted infections (STIs), particularly HIV. In sub-Saharan Africa (SSA), young girls and women accounted for 62% of all new HIV infections in 2023 (UNAIDS, 2024). Despite the availability of other HIV preventative methods, demand for prevention remains stubbornly low in this setting.

Existing economic studies on the demand for new bio-medical prevention methods in low-income settings have focused on men. They show demand to be generally insensitive to information interventions (Godlonton et al., 2016; Thornton, 2008) and price subsidies (Chinkhumba et al., 2014), and only modestly responsive to small financial incentives (Thirumurthy et al., 2014; Thirumurthy et al., 2016). Some of the above have also studied distance and convenience costs of prevention and find limited evidence that this is a barrier amongst men. As far as we are aware, none study agency costs of mitigating risk, which is particularly relevant for women in these settings. Studies on women have focussed on teenagers in schools or public libraries and found positive effects of relative risk information on risk awareness and risky sexual behaviour (Datta et al., 2015; Dupas, 2011; Dupas et al., 2018). However, there is no evidence on whether these positive effects extend to a general population of young adult women who are also at high risk, and importantly whether correcting inaccurate prior beliefs along with information on benefits of prevention leads to an increase in take-up of a preventative health product (Kremer et al., 2019).

To fill this gap, we conducted a randomised-controlled experiment to test the hypothesis that demand for a new (in this setting), free preventative biomedical health product responds to a combination of – adjustment of prior beliefs on relative risks of infection and benefits of prevention and clinic referral enabling access to the product. To our knowledge, this is the first study to test this hypothesis and directly study demand for a new HIV prevention product amongst the general population of young adult women (18-24years) in a low-income, high-risk setting.

Our intervention comprised two core components – an intensive information component and provision of a clinic referral for PrEP initiation. The information component involved a digital self-administered feedback-based ‘quiz’ on relative risks of HIV infection and reduction in risks from HIV testing, condom use and PrEP. At the end of the quiz individuals could optionally provide their phone numbers for a call with a nurse from their local clinic to discuss PrEP. But almost none did. Finally, all participants were provided with a referral letter for their local clinic to initiate PrEP. Control arm participants received no intervention. As part of the experiment, PrEP was made available in participating public clinics in both arms. Our primary outcome is take-up of the product within six months. We also study the effects on risk beliefs and sexual behaviour. A feature of our study is the availability of baseline measures of time preferences, measured with monetarily incentivized economic experiments and a biomarker of baseline risky sexual behaviour. This allows us to evaluate heterogeneity in demand for prevention by present-biased preferences and baseline risk levels.

First, we find that the information component resulted in a large correction of risk beliefs that is sustained over six months. Second, within six months our intervention leads to an increase in demand for the preventative health product. The impact amongst those participating in the intervention is an absolute increase of 5.8 percentage points relative to the control group. Given very low baseline levels of this new technology, this implies a large (>100%) increase relative to the control mean. Within a short period, this is a positive sign and generates hope for the expansion of PrEP. We find this demand is primarily driven by a sub-group of older (20-24 years) women in our sample. Reassuringly, we find no evidence that present-bias or baseline risk levels are a barrier to demand for prevention.

Our study makes important contributions to the literature on demand for prevention. We contribute to the literature studying the role of information interventions in improving demand for prevention. Several studies demonstrate no or small effects of information or education campaigns on prevention use (Dupas, 2009; Godlonton et al., 2016; Kremer & Miguel, 2007; Meredith et al., 2013; Thornton, 2008). While the effect of relative risk information on the usage of preventative health care is unknown (Kremer et al., 2019). The general conclusion from this literature is that ‘light-touch’ information interventions have little to no impact, while intensive information or specific information interventions are more likely to be successful (Dupas & Miguel, 2017). To our knowledge our study is the first to assess the effects of HIV related relative risk information on correction of risk beliefs amongst the general population of adult women and provide evidence on whether this improved understanding results in demand for a new preventative health product.

Preventative interventions for HIV are now free and widely available, however, the non-monetary costs remain high, particularly for women in SSA. Gender and social norms imply women have limited agency to negotiate their own use of a female condom or that their male partners use a condom or take up voluntary medical male circumcision (VMMC) (Duby et al., 2023; Eaton et al., 2003; Seidu et al., 2021). This limited ability to act may lead to overly optimistic beliefs about health risks resulting in underconsumption of prevention (Banerjee & Duflo, 2011; Schwardmann, 2019). Existing studies of non-monetary costs summarised in Dupas and Miguel (2017) have primarily focussed on distance and convenience costs, including for HIV. By focussing on PrEP, a free prevention method, who’s use is promoted globally as being largely within the control of women and providing clinic referrals for PrEP initiation, we increase a woman’s agency to mitigate risk.

Baseline preferences may also affect the demand for preventative health care (Newhouse, 2021). Present biased preferences may cause individuals to procrastinate on engaging in preventive behaviours or place greater weight on the short-term costs of prevention than on the longer-term benefits of avoiding an STI (O’Donoghue & Rabin, 1996). However, the role of present bias in underutilization of prevention in developing countries is not yet clearly established (Dupas & Miguel, 2017). We directly contribute to this literature by assessing present bias as a barrier to demand for prevention (Dupas & Miguel, 2017).

The rest of this paper is organised as follows. Section 2 describes our study setting and the experimental design. Section 3 describes our data and methods. Section 4 describes our findings and Section 5 offers a discussion of results and study conclusions.

## 2. Study setting and experiment design

### 2.1. Study setting

The experiment took place in 6 communities in Manicaland Province, Eastern Zimbabwe. The communities include two peri-urban small towns, three rural communities and one high-density urban suburb. Each community is served by one or more public health facilities that offers HIV services. Baseline data were gathered between July 2018 and December 2019. In all communities baseline data were gathered from all 15-24 year-old women and men as part of a larger study on behavioural drivers of risky sexual behaviour (Thomas et al., 2024). Thus, our experiment participants are a subpopulation of the women in these communities - aged 18-24 years. Baseline data collection activities included a detailed individual socio-behavioural survey and incentivised experimental tasks to measure risk, time, and social preferences. All participants were offered provider-initiated HIV counselling and testing, and dried blood spot (DBS) samples were gathered for laboratory-based testing.

HIV prevalence in 2018-19 was high across study communities with 12.32% of women and 9.81% of men HIV positive. As in previous studies, HIV prevalence rises with age (Figure S1). Amongst all women aged 18-24 years in the study areas, 87.38% had completed secondary education or higher; 58.53% were currently married and 63.15% were unemployed (Panel A of Table S1). 3.00% of the women in this age group were HIV-positive and 29.82% were HSV-2 positive (a well-established proxy of risky sexual behaviour (Baird et al., 2012; Cowan et al., 1994)). 96.03% reported a low chance of becoming HIV positive in the next 12 months. Only 36.50% correctly identified an older man (25-29 years) as being more likely to be HIV-positive than a younger man (15-19 years). Condom use was low with 14.42% reported using a condom in their last sexual encounter (Panel B of Table S1).

### 2.2. Pre-exposure Prophylaxis for HIV (PrEP)-background and availability

PrEP available in SSA at the time of the study, was a combination of emtricitabine/ tenofovir disoproxil fumarate (FTC/TDF) taken orally once-a-day for the prevention of HIV. If taken consistently PrEP has been shown to be >90% effective in preventing HIV (Baeten et al., 2012; Marrazzo et al., 2015; Van Damme et al., 2012). Evidence from clinical trials in SSA indicate low uptake and adherence amongst women are due to underestimation of their risks of infection, and uncertainty and ambivalence about using antiretrovirals for prevention (Corneli et al., 2015; van der Straten et al., 2014).

Since December 2016, Zimbabwe has been implementing a phased national roll-out of PrEP. National guidelines for HIV prevention were updated to include oral PrEP as an additional prevention option for individuals at high risk of HIV (MOHCC, 2017). The definition of high risk includes but is not limited to – HIV-negative partners in a sero-discordant relationship, female sex workers, young women (15-17) selling sex and those in transactional partnerships, married or single people in relationships that put them at risk of HIV infection and people in relationships with partners of unknown HIV status. PrEP had not formally reached public health facilities in study communities prior to the time of our experiment. At the time, it was available to participants in research projects and select clinics funded by international donors. Thus, we collaborated with the Ministry of Health and Child Care to make PrEP available in local public health facilities in both arms prior to the experiment.

As part of the experiment, participants presenting at clinics in intervention and control arms were eligible for PrEP regardless of whether their ‘risk status’ placed them in any of the categories defined above. Study clinics followed national clinical guidelines for PrEP initiation, which include an HIV test, pregnancy test and STI screening as part of the PrEP initiation package. At baseline less than 1% of participants report being on PrEP (Panel B of Table S1). However, most intervention participants – based on analysis of responses to intervention questions - had heard of PrEP, knew how it is taken and its benefits in reducing risk of HIV (Figure S3).

### 2.3. Experiment design

In each community two clusters of villages were created at the start of the study. Within each community one cluster was then randomly allocated to the intervention arm of the experiment while the other was allocated to the control arm. Thus with 6 clusters per arm, assuming a mean cluster size of 54 individuals, intraclass correlation coefficient (ICC) of 0.01 and baseline PrEP usage of 0.05%, the experiment had >80% power to detect an increase of ∼5 percentage points in PrEP take up. ^1^

Following baseline data collection and HIV testing, a random sample of eligible women were invited to participate in the experiment. Women in the study areas were considered eligible to participate in the experiment if they were aged between 18–24 years and resident in the experiment clusters. Women were excluded if they had tested positive for HIV at baseline or self-reported taking PrEP at baseline. Women in intervention clusters received the intervention, while those in control clusters received no intervention.

Eligible participants in intervention communities were invited to a central location local in the cluster (school, hall or tent erected for study purposes etc). On arrival, participants were informed they were about to take part in a study and informed consent was sought. The information component of the intervention was delivered individually to each participant via a digital tablet in the local language Shona. Experimenters provided participants with training on how to use the tablet. The entire session took a participant approximately 30 minutes to complete.

The aim of the feedback-based information component was twofold – first, to correct prior misbeliefs about risks of HIV by partner age, and second, to increase awareness and understanding of risk reduction strategies including HIV testing, condom use and PrEP. This component of our intervention builds on Dupas et al (2018, 2011) and Datta et al (2015) – who focused on information regarding risk from older partners, but our intervention is more individualised and extends to risk reduction from various prevention methods. The information was delivered as an interactive, visual scenario based ‘quiz’. The ‘quiz’ begins by asking the respondent to enter their current age. In each scenario the protagonist is a woman who takes on the age of the respondent. Participants then respond to up to 10 rounds of a quiz, where in each round, they are asked to choose which one of two hypothetical characters was more likely to have HIV. Both the age and sex of the individuals was randomly varied and in cases where atleast one of the individuals was a woman, the presented character took on the age provided by the respondent. Participants received immediate feedback on their choices and were presented with visual representations of age-and-sex-specific HIV prevalence amongst individuals represented in that round of the ‘quiz’. The data were calibrated to local HIV-prevalence estimates. If participants made a wrong choice, following feedback, they were presented with another pair of characters of the same sex and age as the original question to reinforce the information. The quiz then extends to a series of scenarios between the protagonist and a male partner approximately 5 years older and covers risks of HIV from when they do not know their HIV status, to potential risk reduction for the protagonist from condom use. Finally, the scenarios introduce the protagonist to PrEP, its benefits, usability and local availability. All participants were then provided with a referral letter to a participating local clinic where they could initiate PrEP. At the end of the session participants could optionally provide their phone numbers for a call with a nurse to discuss PrEP. But almost none chose to do so. In the very few cases who did, nurses did not actually follow up due to their busy schedules.

## 3. Data and Empirical Strategy

### 3.1. Outcomes and hypotheses

Our primary outcome is the proportion of women taking up PrEP within six months, gathered from clinic data and triangulated with self-reports and biomarkers of PrEP presence from DBS. In the latter cases, if an individual self-reported being on PrEP during the follow-up survey, but was not identified from clinic records, then they were requested to provide a DBS which was used to confirm presence of PrEP in the blood sample.

We also study three self-reported secondary outcomes measured six months after the intervention. The first two of these measure risk beliefs and cover knowledge of HIV risks from partnerships with older men and benefits of different prevention options, especially PrEP. These outcomes aim to evaluate whether women update their prior beliefs based on the information component of the intervention. The first measure asked participants to identify which of a 15-19-year-old man or a 25-29-year-old man was more likely to be HIV positive, while the second, asked participants to identify which of a 15-19-year-old woman and a 25-29-year-old man was more likely to be HIV positive. We would expect intervention participants to be more likely to identify older men as being more likely to be HIV positive. By measuring outcomes six months after the intervention, we also evaluate retention of adjusted risk beliefs for a longer period than Datta et al (2015) who measure outcomes immediately after the intervention and in a non-random subsample at three months.

Our third self-reported outcome-proportion of women who report condom use in their last sexual encounter is an indicator of risky sexual behaviour. The effect of introducing information on relative risks of HIV on behaviour depends on how individuals respond to the information. In particular, whether they revise their beliefs of personal risk upwards or downwards, resulting in opposing behavioural responses. If they update their risk beliefs upwards, the resulting change is an expected reduction in risky behaviours, as demonstrated by Dupas et al. (2018, 2011) and Godlonton et al (2016). On the other hand, a reduction in perceived risk may result in either no change, or an increase in risky behaviours – reflecting risk compensation. Similarly, the introduction of a risk reducing health product like PrEP, while strictly decreasing personal risk, in practice, the net impact is ambiguous as risk-compensating behaviours may offset the benefits of the product. Thus, our behavioural outcome allows us to evaluate whether the intervention had any negative spillovers. While we would expect the information provided to increase condom use, it is possible that the availability of PrEP encourages substitution away from condom use. Thus, the direction of expected impact of the intervention on risky behaviours is ambiguous. We emphasize though, that we are only powered to detect with precision moderate to large changes in condom use i.e with an ICC of 0.023, the study has 80% power to detect a 10p.p decrease in condom use.

### 3.2. Measurement of present bias and risk aversion

Time preferences were measured using monetarily incentivised Money Earlier or Later (MEL) experiments (Cohen et al., 2020) with two multiple price lists (MPL) where participants were asked to make a series of choices between a smaller-sooner (SS) ($20) offered right away/in one-month and a larger-later (LL) ($20+X) reward available in 3/4 months (see Thomas et al (2024) for details). Using the last question in each MPL before a participant switched from a SS to LL reward, we derived the implied discount factor (δ_1,4_ and δ_0,3_) and classified an individual as present-biased if δ_0,3_ < δ_1,4_. We measured risk preferences using the “multiple lotteries method” (Binswanger, 1981; Castillo et al., 2011; Dave et al., 2010; Eckel & Grossman, 2002). Participants had to choose one of six gambles to play, with the payoff determined by a coin toss. Each gamble involved a 50/50 chance of a low or high payoff and one safe option. We use the midpoint of the derived range of the coefficient of risk aversion ‘γ’ as an individual’s implied measure of risk aversion.

### 3.3. Validation of the Experiment Design

The randomized design of our experiment offers a straightforward source of identification. The random assignment of clusters of villages to treatment and control groups ensures that on average, women in the intervention group will be similar in socio-economic and behavioural characteristics to those in the control group, except for their exposure to the intervention. Table 1 shows summary statistics of baseline characteristics and outcomes by treatment groups. All but one factor has a normalised difference under 0.13 indicating good balance between the treatment groups (Imbens, 2015). The two groups only differ in the proportion reporting the category ‘Other’ as their economic sector (normalised difference 0.14). Despite a normalised difference under 0.13, it would also appear the treatment group has fewer currently married women (54%) than the control group (63%). In all our models we control for baseline measures of age, education and marital status.

**Table 1:** Characteristics and outcomes by treatment clusters and balance checks.

|  | **Control Clusters**<br>**N=462** | **Intervention**<br>**Clusters**<br>**N=505** | **Normalised**<br>**difference** |
| --- | --- | --- | --- |
|  | Mean [SD] / % | Mean [SD] / % | (C-T) |
| **Panel A: Characteristics** |  |  |  |
| **Age in years** | 20.98 | 20.87 | 0.05 |
|  | [2.04] | [2.09] |  |
| **Education** |  |  |  |
| None/primary | 0.136 | 0.117 | 0.059 |
| Secondary/higher | 0.864 | 0.883 | 0.059 |
| **Marital status** |  |  |  |
| Never married | 0.297 | 0.349 | 0.111 |
| Currently married | 0.617 | 0.556 | 0.123 |
| Divorced/separated | 0.082 | 0.091 | 0.031 |
| Widowed | 0.004 | 0.004 | 0.006 |
| **Economic sector** |  |  |  |
| Estates: tea, coffee, forestry | 0.035 | 0.022 | 0.078 |
| Retail or professional services | 0.028 | 0.026 | 0.007 |
| Informal sector | 0.070 | 0.055 | 0.062 |
| Student | 0.145 | 0.150 | 0.013 |
| Unemployed: excl. agriculture | 0.642 | 0.623 | 0.040 |
| Other | 0.080 | 0.124 | 0.144 |
| **Preferences** |  |  |  |
| Risk aversion | 4.15 | 4.23 | 0.041 |
|  | [2.07] | [2.04] |  |
| Mean discount factor | 0.653 | 0.667 | 0.076 |
|  | [0.192] | [0.198] |  |
| Present biased | 0.203 | 0.245 | 0.101 |
| **HSV-2 status at baseline** |  |  |  |
| Negative | 0.605 | 0.592 | 0.026 |
| Positive | 0.299 | 0.297 | 0.005 |
| Equivocal | 0.096 | 0.111 | 0.049 |
| **HIV status at baseline** |  |  |  |
| Positive | 0.029 | 0.025 | 0.024 |
| **Perceived chance of being infected with HIV in the next 12 months** |  |  |  |
| High | 0.008 | 0.011 | 0.037 |
| Moderate | 0.036 | 0.025 | 0.064 |
| Low | 0.956 | 0.964 | 0.038 |
| **Age at first sex** | 17.93 | 17.90 | 0.017 |
|  | [1.87] | [2.02] |  |
| ***Panel B: Outcomes*** |  |  |  |
| Currently taking PrEP | 0.002 | 0.008 | 0.081 |
| Correctly identified 25-29 year-old man as higher risk of HIV compared to a man 15-19 years old | 0.361 | 0.368 | 0.014 |
| Correctly identified 25-29 year-old man as higher risk of HIV compared to a woman 15-19 years old | 0.191 | 0.202 | 0.027 |
| Condom use at last sex | 0.125 | 0.162 | 0.107 |

### 3.4. Participation and Attrition

Following HIV testing at baseline, based on experiment eligibility defined earlier, a random sample of eligible women were selected to participate in the experiment (N=749). The approach to include a random sample of eligible baseline participants was an ethics committee requirement to minimise the likelihood of unintentional HIV status disclosure within communities based on experiment eligibility. If all HIV-negative women within a community were invited to participate in a prevention intervention, then it would potentially become known within a community that those not invited were HIV-positive. In the treatment clusters, randomly selected participants (N=380) were invited to a central location local to the cluster, where the intervention was delivered. Of those invited in the treatment clusters 76% took up the invitation and participated in the intervention. In the control clusters no intervention was delivered, and participants were followed up six months later (N=369). To assess characteristics associated with participation in the intervention, we regressed a binary indicator of participation (1=treatment arm complier, 0=control arm or non-complier from treatment arm) on a range of baseline socio-economic characteristics. We do not find evidence of selection into participation based on observable characteristics (Table S2). In our analysis we estimate local average treatment effects (LATE) to recover unbiased treatment effects for the participants.

After six months, all invited participants were included in the follow-up activities – regardless of whether they participated in the intervention or not. Follow-up activities included repeating the socio-behavioural survey along with DBS collection amongst those reporting being on PrEP. 14% (N=52) of women in the treatment arm and 15% (N=56) of women in the control arm were lost-to-follow-up (Figure S2). This attrition rate is not surprising amongst young people in general population cohorts, particularly as women in this age group are likely to move away either due to marriage or to find work. At approximately 85% our retention rate is similar or higher than other recent studies in sub-Saharan Africa. For example, the recently concluded PopART HIV Study in Zambia and South Africa had an annual retention rate of 72% in its population cohort (Hayes et al., 2019). We find no evidence of differential attrition for any outcome (Table S3). In general, we find our control and treatment follow-up samples are similar across a range of characteristics with normalised differences less than 0.13 (Table S4). They only differ in marital status - with 54% of the treatment sample being married, compared to 63% in the control sample. As mentioned earlier, in all our models we control for age, education and marital status to account for any small imbalances.

### 3.5. Empirical Strategy

Using instrumental variable analysis, we estimate local average treatment effects (LATE) which provide the causal effect of participating on compliers (Angrist et al., 1996). We estimate LATEs using two-stage least squares (2SLS) with random assignment as an instrument for intervention participation. All models include community (randomization cluster) fixed effects, age, education and marital status as covariates. To help assess the magnitude of our estimates, we report average outcomes for the control group women. We report heteroskedasticity-robust standard errors clustered at the randomisation cluster level.^2^

To assess robustness, we re-estimate our models with our main set of covariates and baseline outcomes, and with a full set of covariates and baseline outcomes. In the latter case, in addition to age, education and marital status, we control for perceived risk of infection in the next 12 months, mean discount factor across the MPLs, present bias, risk aversion and a binary indicator of whether they were a consistent respondent in the time preference exercise. We also assess robustness to specifications without clustering standard errors and estimation with wild restricted efficient bootstrap standard errors. The latter is an extension of the wild cluster bootstrap approach for models with few clusters, to linear regression models estimated with instrumental variables (Davidson & MacKinnon, 2010).

Across our outcomes, we estimate heterogeneity by baseline - age-group (18-19-year-olds and 20-24-year-olds), present biased preferences and risk levels. Given age of reported sexual debut in our sample is ∼18 years, we would expect older women to be more sexually active and therefore more at risk either through number of partners or marriage. We test for heterogeneity by interacting the treatment indicator with either a binary measure of age-group (18–19-year-olds =0/20-24-year-olds=1) or present bias (no present bias=0, present bias=1). In the models for heterogeneity by present bias, in addition to age, education and marital status, we control for risk aversion, the mean discount factor across the two MPLs and a binary indicator of whether the individual was a consistent respondent in the time preference exercise.

We follow the economics literature and study how information interacts with baseline risk (Boozer & Philipson, 2000; Godlonton et al., 2016; Gong, 2015).^3^ Women at high-risk of infection are most likely to benefit from PrEP and the intervention would be most successful in reducing HIV incidence if high-risk women took up PrEP. Holding other factors constant, high-risk women receiving information of their risky status via the intervention would be expected to take up PrEP. However, high-risk women may either have greater willingness to bear risk or low agency to use prevention, limiting take-up in this group. On the other hand, women who ex ante are low-risk or practice safe sex, on receiving information of their low-risk status, may by more likely to take-up PrEP when offered. We interact our treatment indicator with a biomarker of HSV-2 status. HSV-2 status is widely used proxy measure of risky sexual behaviour (Baird et al., 2012; Cowan et al., 1994; Thomas et al., 2024). Being a biomarker, HSV-2 infection status is an accurate composite measure of exposure to risk, capturing direct risk from an individual’s behaviour, as well as being at risk via a partner’s sexual behaviour. As shown in Table S1, at baseline 29.82% of women in our young study sample were already HSV-2 positive. Since women were not informed of their HSV-2 status following laboratory testing, they may not be aware of their high-risk status prior to the intervention.

## 4. Results

### 4.1. Main effects

In Table 2, we present estimates from our 2SLS models. We find the intervention increased PrEP take-up in the treatment group by 5.8 percentage points (p.p) (p<0.05). This implies a modest absolute increase in the take-up of PrEP, but a large relative increase from the low control mean of 0.015.

**Table 2:**
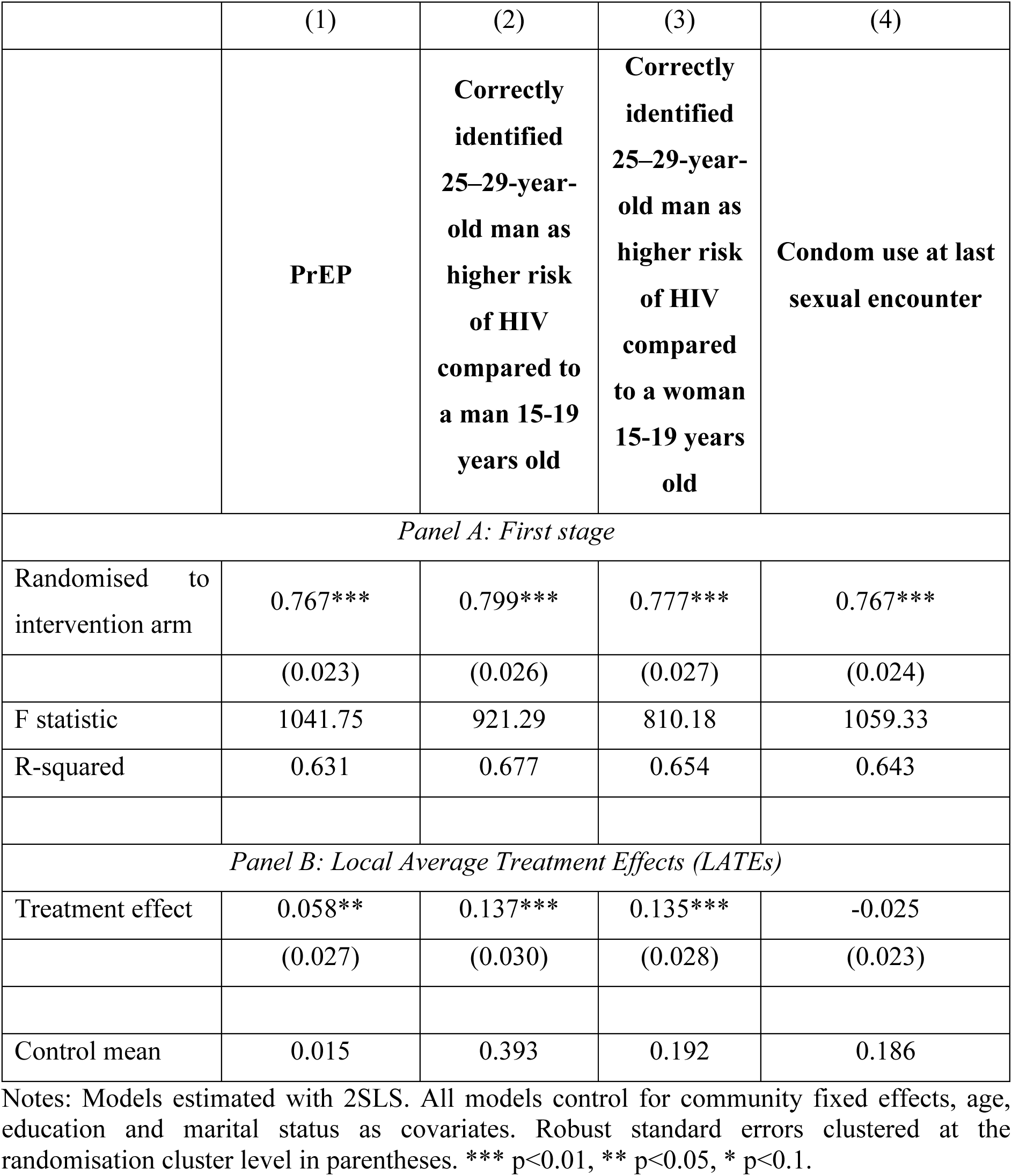
Impact of the intervention. Notes: Models estimated with 2SLS. All models control for community fixed effects, age, education and marital status as covariates. Robust standard errors clustered at the randomisation cluster level in parentheses. *** p<0.01, ** p<0.05, * p<0.1.

Our baseline data indicates that most individuals held inaccurate beliefs about HIV risk from older partners, with only ∼36% likely to identify a 25-29-year-old-man being more likely to be infected than a 15-19-year-old-man (See Table 1). Evidence from the follow-up survey suggests that respondents in the treatment group updated their beliefs. We find a statistically significant and large increase of 13.7 p.p (p<0.01) in the proportion of treatment group women correctly identifying a 25-29-year-old man was more likely to be HIV positive than a 15-19-year-old man (Table 2 column 2). We also find a 13.5 p.p (p<0.01) increase in the proportion accurately identifying a 25-29-year-old man as being more likely to be HIV positive compared to a 15-19-year-old woman (column 3). These estimates imply a 35% and 70% increase relative to the control mean.

For the risky behaviour outcome - condom use at last sexual encounter, the direction of the coefficient indicates a decline in condom use by 2.5 p.p (13%) which is not statistically significant. As described earlier, the study was powered to detect with precision moderate to large changes in this outcome ∼10p.p or greater. However, our negative point estimate is suggestive of risk compensation.

The results presented above are robust to the inclusion of baseline outcomes and a wider set of covariates (Table S6). Importantly, in the model of condom use at last sexual encounter, controlling for baseline condom use improves the magnitude and precision of the estimates (−0.047, p<0.10). This is not surprising since there was a slight imbalance for this outcome in the follow-up sample (Table S4). The estimates are also robust to specifications without clustering standard errors and wild restricted bootstrap standard errors (Table S7).

### 4.2. Heterogeneity in local average treatment effects

In Table 3, we present coefficients for the interaction of the treatment indicator with each binary covariate (age group, present bias and HSV-2 status) and p-values from a joint test of equality of the treatment effect between covariate sub-groups. Considering age-group differences in treatment effects, we find PrEP take up increased amongst older women (20-24years) by 7.2 p.p (p<0.05). In contrast, take up amongst 18-19-year-olds was negligible (0.007 p.p) and not statistically significant (column (1)). The test of equality rejects the null (p<0.05) that the treatment effect is similar in the two age groups, confirming heterogeneity in the take-up of PrEP by age-group.

**Table 3:**
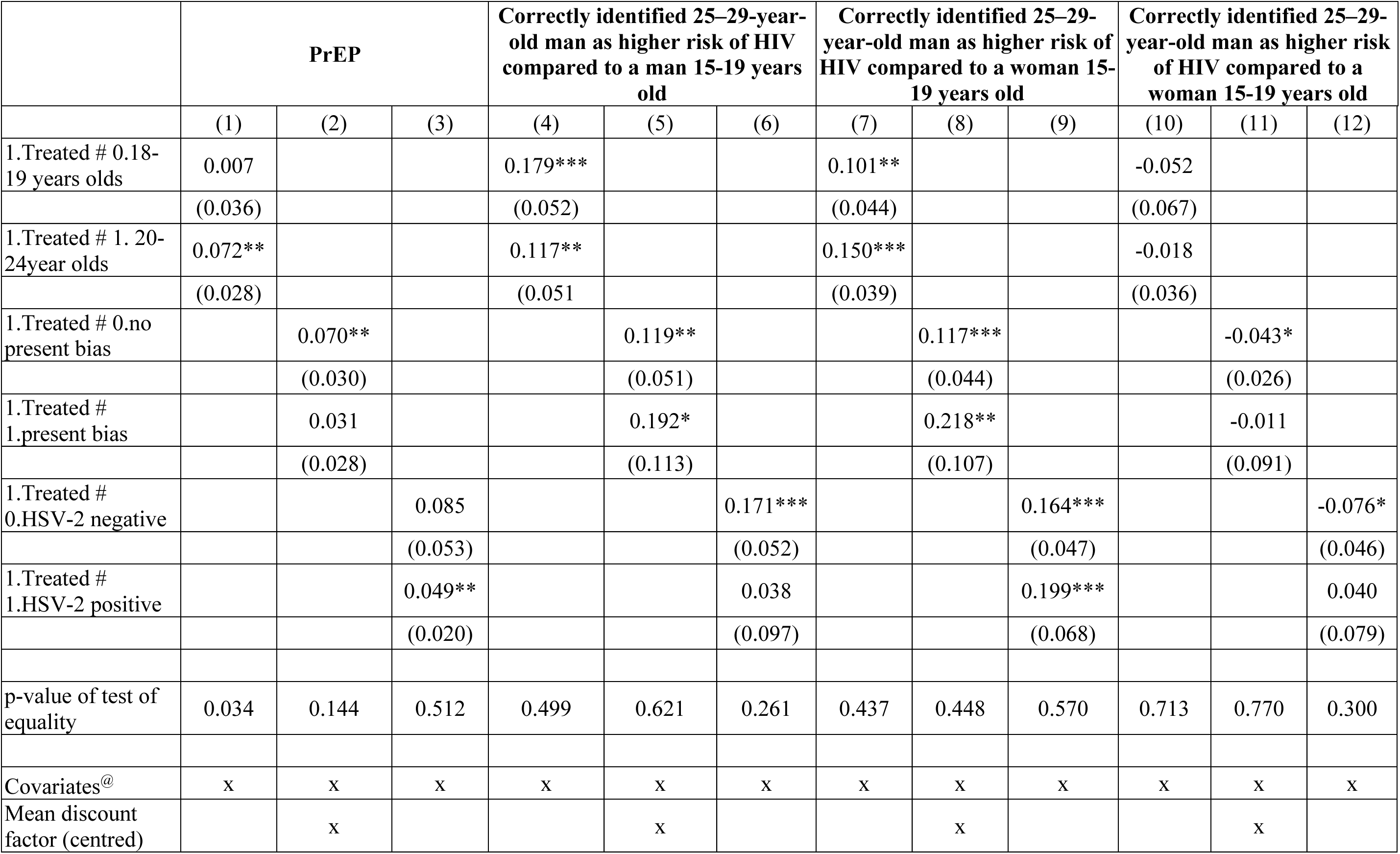
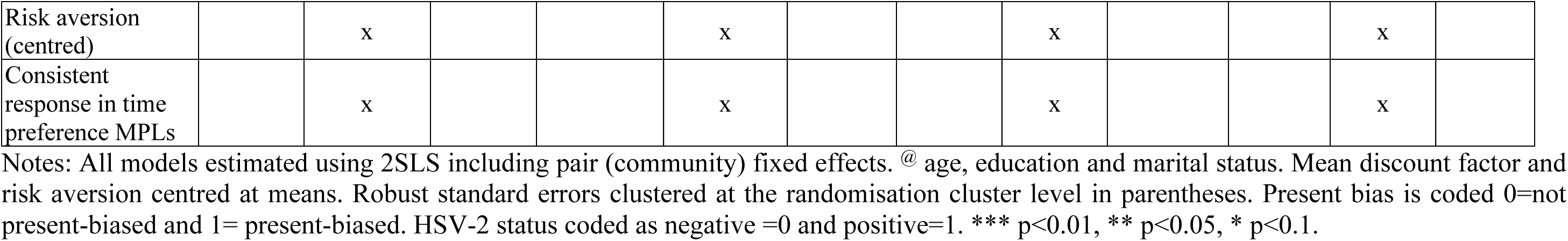
Heterogeneity by baseline age, present-biased preferences and risky behaviour. Notes: All models estimated using 2SLS including pair (community) fixed effects. @ age, education and marital status. Mean discount factor and risk aversion centred at means. Robust standard errors clustered at the randomisation cluster level in parentheses. Present bias is coded 0=not present-biased and 1= present-biased. HSV-2 status coded as negative =0 and positive=1. *** p<0.01, ** p<0.05, * p<0.1.

|  | PrEP |  |  | Correctly identified 25–29-year-old man as higher risk of HIV compared to a man 15-19 years old |  |  | Correctly identified 25–29-year-old man as higher risk of HIV compared to a woman 15-19 years old |  |  | Correctly identified 25–29-year-old man as higher risk of HIV compared to a woman 15-19 years old |  |  |
| --- | --- | --- | --- | --- | --- | --- | --- | --- | --- | --- | --- | --- |
|  | (1) | (2) | (3) | (4) | (5) | (6) | (7) | (8) | (9) | (10) | (11) | (12) |
| 1.Treated # 0.18-19 years olds | 0.007 |  |  | 0.179*** |  |  | 0.101** |  |  | -0.052 |  |  |
|  | (0.036) |  |  | (0.052) |  |  | (0.044) |  |  | (0.067) |  |  |
| 1.Treated # 1. 20-24year olds | 0.072** |  |  | 0.117** |  |  | 0.150*** |  |  | -0.018 |  |  |
|  | (0.028) |  |  | (0.051) |  |  | (0.039) |  |  | (0.036) |  |  |
| 1.Treated # 0.no present bias |  | 0.070** |  |  | 0.119** |  |  | 0.117*** |  |  | -0.043* |  |
|  |  | (0.030) |  |  | (0.051) |  |  | (0.044) |  |  | (0.026) |  |
| 1.Treated # 1.present bias |  | 0.031 |  |  | 0.192* |  |  | 0.218** |  |  | -0.011 |  |
|  |  | (0.028) |  |  | (0.113) |  |  | (0.107) |  |  | (0.091) |  |
| 1.Treated # 0.HSV-2 negative |  |  | 0.085 |  |  | 0.171*** |  |  | 0.164*** |  |  | -0.076* |
|  |  |  | (0.053) |  |  | (0.052) |  |  | (0.047) |  |  | (0.046) |
| 1.Treated # 1.HSV-2 positive |  |  | 0.049** |  |  | 0.038 |  |  | 0.199*** |  |  | 0.040 |
|  |  |  | (0.020) |  |  | (0.097) |  |  | (0.068) |  |  | (0.079) |
| p-value of test of equality | 0.034 | 0.144 | 0.512 | 0.499 | 0.621 | 0.261 | 0.437 | 0.448 | 0.570 | 0.713 | 0.770 | 0.300 |
| Covariates<sup>@</sup> | x | x | x | x | x | x | x | x | x | x | x | x |
| Mean discount factor (centred) |  | x |  |  | x |  |  | x |  |  | x |  |
| Risk aversion<br>(centred) |  | x |  |  | x |  |  | x |  |  | x |  |
| Consistent<br>response in time<br>preference MPLs |  | x |  |  | x |  |  | x |  |  | x |  |

In column (2), results of heterogeneity by an indicator of present bias indicates that amongst time-consistent individuals, the intervention led to a 7.0 p.p (p<0.05) increase in take-up of PrEP, while amongst present-biased individuals there was a statistically insignificant 3.1 p.p increase. The joint test of equality fails to reject a null of equality in treatment effects between these two groups.

In column (3) we find PrEP take-up increased in both baseline risk groups. Amongst high-risk (HSV-2 positive) women the intervention led to a 4.9 p.p (p<0.05) increase and amongst low-risk (HSV2-negative) women we find a larger 8.5 p.p increase, but this is not statistically significant. This indicates the intervention was successful in increasing take up amongst those most likely to benefit from the intervention (high-risk) and as expected amongst low-risk individuals. However, considering the statistically in-significant joint test of equality between the groups, we conclude there is no heterogeneity by baseline risk group.

Columns (4)-(9) of Table 3 show heterogeneity analysis for risk beliefs. For both outcomes, across age groups, preferences and risk levels we find that the intervention led to a statistically significant and large correction of risk beliefs. We do not find evidence of heterogeneity by any of these covariates. Overall, we conclude that the intervention successfully updated risk beliefs across our sample of women.

Columns (10)-(12) present results for our sexual behaviour outcome, condom use. While we do not see statistically significant heterogeneity across groups, the magnitude and direction of estimates present interesting findings. The intervention appears to have generally resulted in a large decrease in condom use across age-groups, time preference groups and amongst low-risk individuals.

## 5. Discussion and conclusions

We find that intensive information on relative risks and prevention benefits, combined with a clinic referral increases demand for a new preventative health product. Considering low baseline levels of usage, our absolute increase of 5.8 p.p reflects a large (>100%) relative increase in take-up of PrEP. This demand appears to be driven by older women (20-24 years-old). We find no evidence of heterogeneity by present-biased preferences or baseline risk levels. The relative risk information component resulted in a large correction of prior risk beliefs of HIV infection which appears to have been sustained over six months. We find indicative evidence of risk compensation resulting from a reduction in condom use following the intervention.

Our finding of increased accuracy of risk beliefs is consistent with previous literature showing targeted or intensive information can have large effects on knowledge of risks and health outcomes. Datta et al (2015), which is closely related to the feedback-based age-disparate risk information component delivered, but focused on a sample of teenagers recruited at a public library, find an 18p.p. increase in the proportion correctly identifying an older man, compared to a younger man as being more likely to be HIV-positive, immediately after the intervention. At three months, they find no significant effect. In contrast, to their latter finding, we show that the effect on updating prior beliefs is sustained over six months in our general population sample of adult women. In a recent paper in Malawi studying adults 45 years and over, Cianco et al (2024) find a video information intervention with visual statistical information on survival chances, resulted in a 6.1% increase in subjective expectation of population survival and 6.5% increase in subjective expectation of contracting HIV from multiple partners after one year.

An important policy question that remains unanswered in the literature, is the extent to which updating prior beliefs on risks and benefits of prevention results in take-up of prevention (Kremer et al., 2019). Three studies closely related to ours in terms of the information conveyed and targeting of prior beliefs - Dupas et al (2011; 2018) and Cianco et al (2024), study risky behaviours. They find large reductions in teen pregnancy incidence (28% by (Dupas, 2011)) and a 19% reduction in the predicted probability of not using a condom with multiple partners (Ciancio et al., 2024). However, none of these studies directly considered take-up of a preventative health product as we do. Datta et al (2015) also only consider knowledge outcomes. Our finding of an increase in take-up shows that correction of prior beliefs can lead to an increase in demand for a new prevention product.

Existing evidence on the effects of information or non-price mechanisms on demand for free preventative health products have found either no effects or smaller effects than ours. In terms of magnitude, the 5.8 p.p increase in take-up within six months in our study, reflects a large increase from very low baseline levels. This is a positive sign within the short follow-up time especially since PrEP was a new bio-medical intervention in our study setting. Other economic studies that have considered new bio-medical HIV prevention methods, have found much smaller results for one-of prevention interventions. Thirumurthy et al. (2014) find small financial incentives lead to 5 p.p (3.1%) uptake in VMMC within 2 months. Chinkhumba et al. (2014) find information on benefits of VMMC increased demand by 66% (1.4p.p), but overall rate of circumcision was low with only 3.1% of those offered free VMMC taking up the procedure. Godlonton et al. (2016), find no statistically significant take-up of VMMC one year after participants were provided information on the benefits of VMMC. Considering non-bio-medical preventative interventions, Banerjee et al. (2017) find that a movie intervention promoting the benefits of double fortified salt (DFS) to prevent anaemia, led to a large 5.5 p.p (57%) higher use of DFS 7-16 months after the intervention. On the other hand, several studies find small or negligible impact of information on demand for preventative health products (Ashraf et al., 2013; Kremer & Miguel, 2007; Meredith et al., 2013).

Over the last few years there has been growing interest in analysing behavioural responses to interventions. The debate on the extent to which present-biased preferences are a barrier to demand for preventative healthcare in developing countries remains unsettled (Dupas & Miguel, 2017; Kremer & Glennerster, 2012; Kremer et al., 2019). Our finding that demand did not differ by present-biased/time consistent preferences makes an important contribution to this debate, highlighting that present-bias may not be a barrier to take-up of prevention.

Reassuringly we find that ex ante high-risk women when made aware of risks and benefits of prevention through the intervention demand PrEP, just as low-risk women. It might seem unusual that we observe take-up amongst women who are at high-and-low-risk regardless of their time preference. An explanation for this is, women in our settings are at risk not just from their own preferences and actions (e.g due to present bias or inherent willingness to bear risk) but are also placed at risk by their partners due to their low negotiating power for the use of prevention within partnerships. HSV-2 status as a biomarker captures both sources of risk. Thus, it is plausible that high risk women regardless of their time preferences demand prevention when made aware of their risk through the intervention.

The modest absolute increase we see in our study within six months is likely due to other non-monetary costs that our intervention did not address. While PrEP has been promoted globally as a method that empowers women by being within their control, access alone does not ensure agency to mitigate risk. In a qualitative study that accompanied our experiment, young women highlighted several limitations to their autonomy to initiate and use PrEP (Skovdal et al., 2022). These included women’s own perceptions that they would require their partner’s permission to use PrEP, fear of being spotted at health clinics for accessing PrEP and stigma around PrEP use, which include being perceived as being HIV positive or a sex worker, and the lack of privacy to consistently take PrEP pills without discovery.

A key concern around new preventative health products has been whether risk compensation would offset the positive technological benefits. For HIV and sexual behaviour, such concerns were expressed when VMMC was first introduced as a HIV prevention strategy. However most empirical studies find limited evidence of risk compensation i.e. that VMMC does not lead to riskier sex amongst circumcised men (Godlonton et al., 2016; Wilson et al., 2014). In the case of PrEP, several public health and bio-medical studies in high- and low-income settings have raised similar concerns. Thus far there is no evidence that risk compensation leads to higher rates of HIV infection amongst PrEP users. However, the concern is also that risk compensation may lead to other negative outcomes such as unwanted pregnancies and STIs (Marcus et al., 2019). We find evidence that condom use decreased by ∼13% following the intervention.

Our study shows that demand amongst young adult women for a new preventative health product responds to intensive information on risks and benefits of prevention, when coupled with free access to the product at public clinics.

## Funding

National Institutes of Mental Health, USA grant R01MH114562-01. Bill and Melinda Gates Foundation OPP1161471. Medical Research Council (MRC), Centre for Global Infectious Disease Analysis funding from the UK Medical Research Council and Department for International Development MR/R015600/1.

## Ethical approval

The study has obtained ethical approvals from the Medical Research Council of Zimbabwe (reference number: MRCZ/A/2243), the Biomedical Research and Training Institute, Zimbabwe Institutional Review Board (reference number: AP140/2017) and the Imperial College London Research Ethics Committee (reference number: 17IC4160).

## Data Availability

All data produced in the present study are available upon reasonable request to the authors

## Supplementary Material

**Figure S 1:**
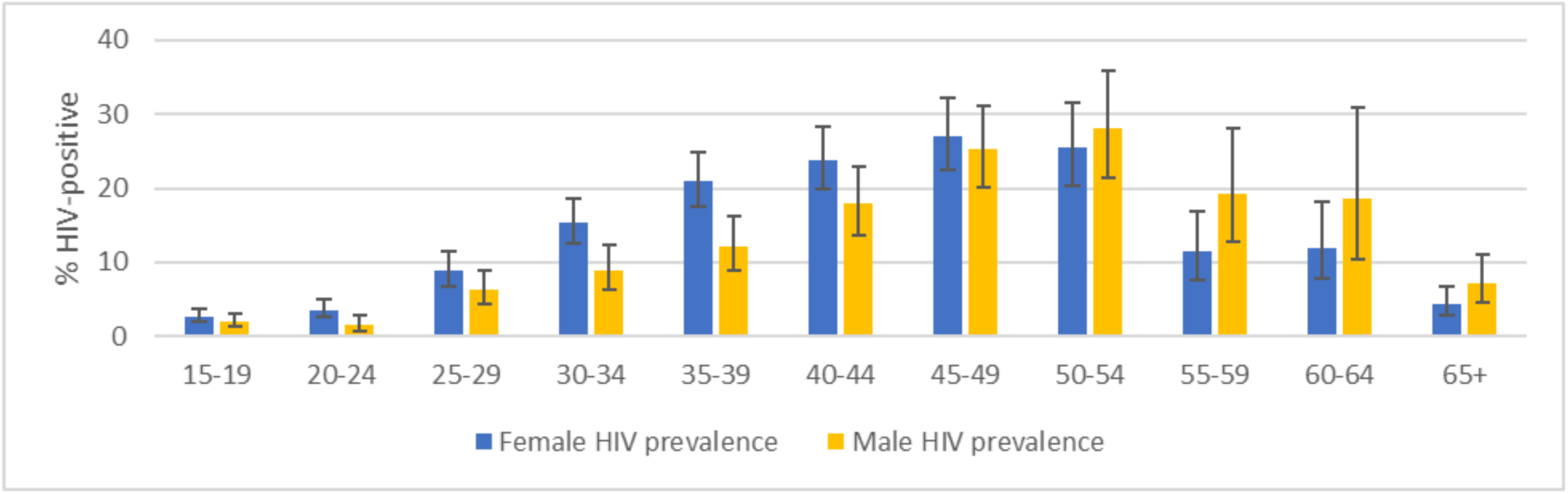
All-age HIV prevalence

**Figure S 2:**
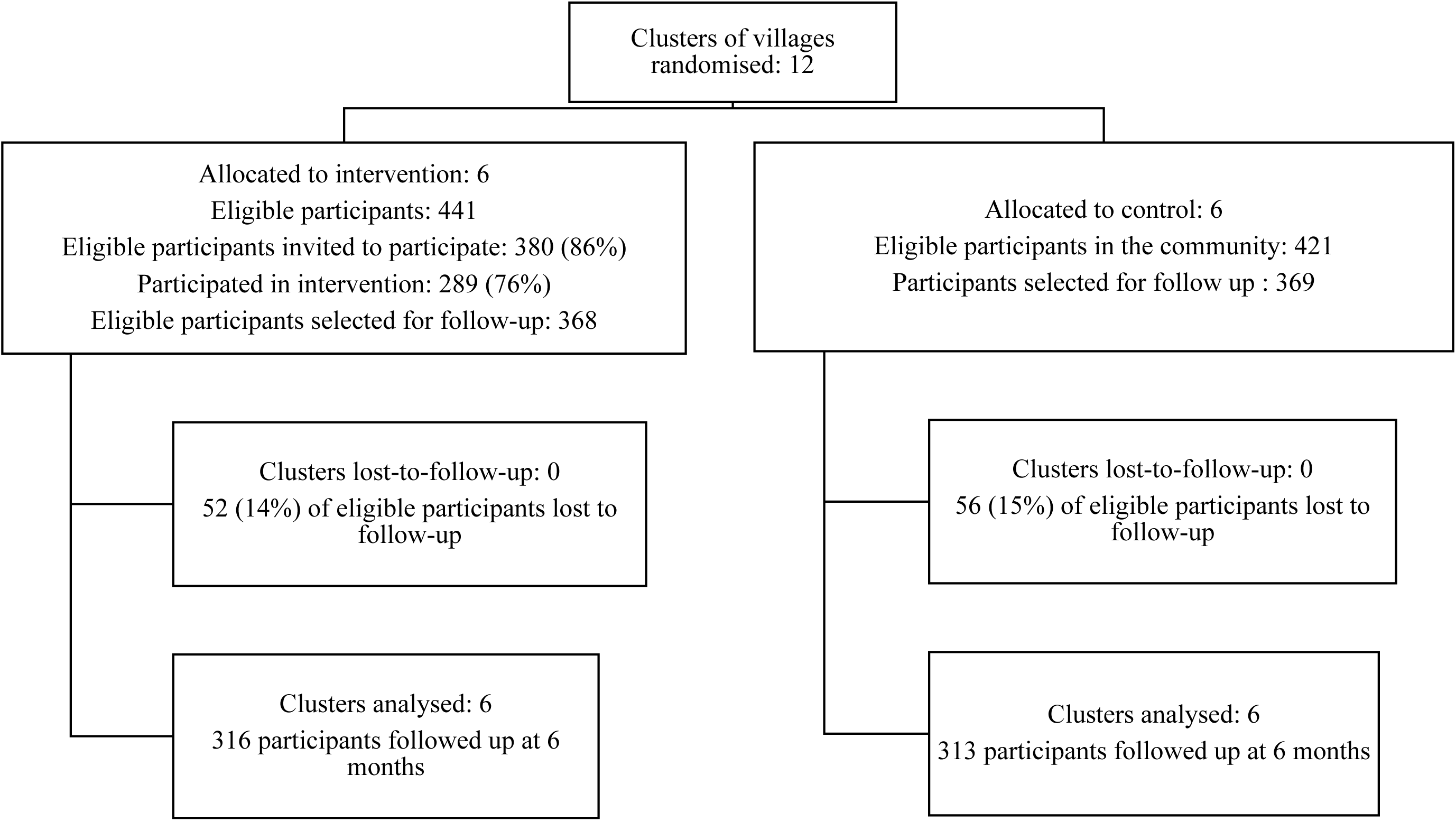
Experiment design, participation and attrition

**Figure S 3:**
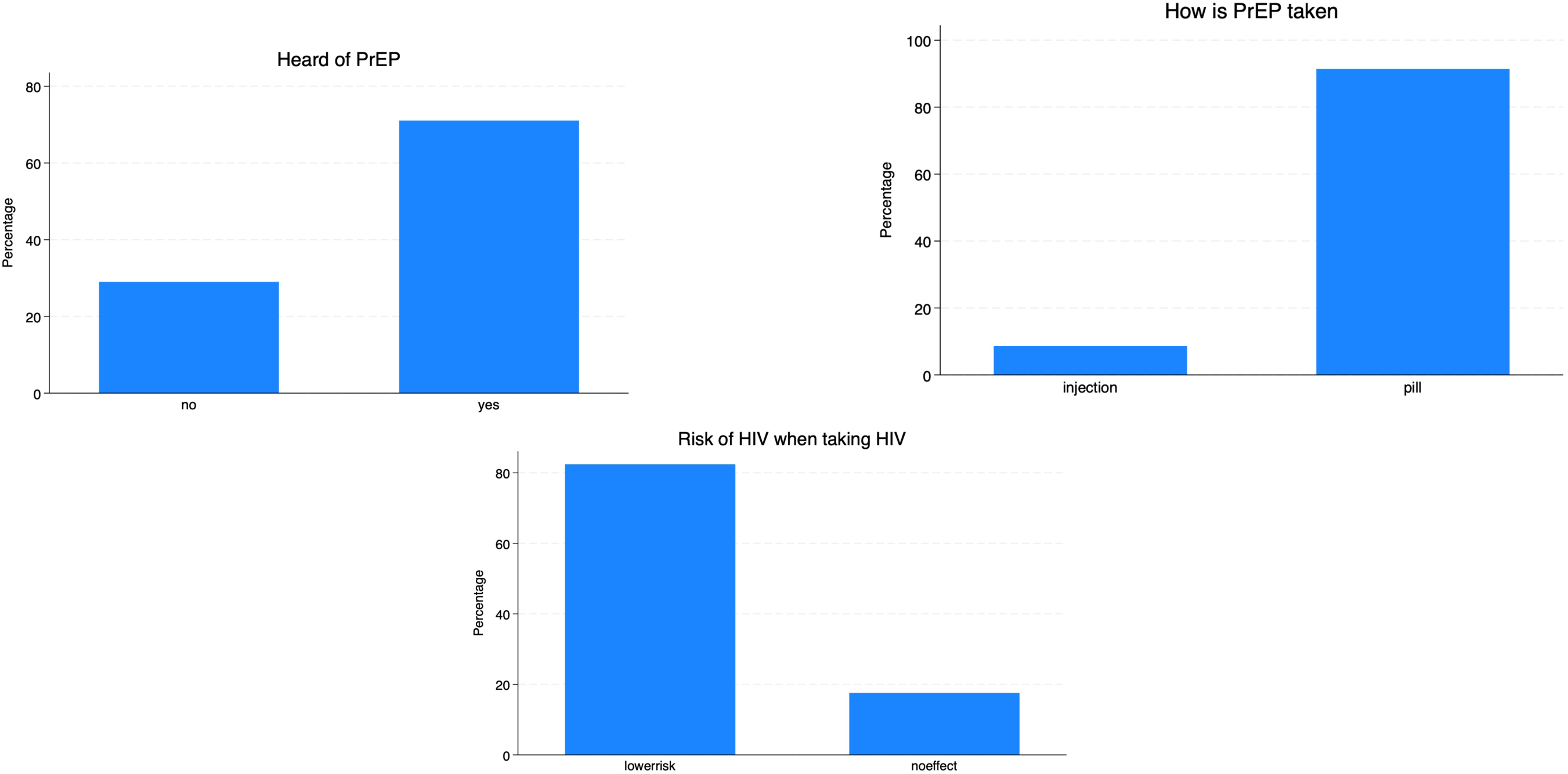
Awareness of PrEP – responses from the information intervention

---

**Table S 1:** Summary statistics on socio-economic characteristics and sexual behaviour among women 18-24 years in Study Area Notes: Dependent variable is a binary indicator equal to 1 if a treatment arm individual participated in the intervention, and equal to 0 for individuals randomised to the control arm or non-compliers in the treatment arm. Estimates obtained through OLS regressions. Robust standard errors clustered at the randomisation cluster level in parentheses. *** p<0.01, ** p<0.05, * p<0.1.

|  | **N** | **Mean**<br>**[SD]/%** |
| --- | --- | --- |
| *Panel A: Socio-economic characteristics* |  |  |
| **Age in years** | 967 | 20.92 |
|  |  | [2.06] |
| **Education** |  |  |
| None/primary | 967 | 12.62% |
| Secondary/higher | 967 | 87.38% |
| **Marital status** |  |  |
| Never married | 967 | 32.37% |
| Currently married | 967 | 58.53% |
| Divorced/separated | 967 | 8.69% |
| Widowed | 967 | 0.41% |
| **Economic sector** |  |  |
| Estates: tea, coffee, forestry | 852 | 2.82% |
| Retail or professional services | 852 | 2.70% |
| Informal sector | 852 | 6.22% |
| Student | 852 | 14.79% |
| Unemployed: excl. agriculture | 852 | 63.15% |
| Other | 852 | 10.33% |
| **Preferences** |  |  |
| Mean discount factor | 852 | 0.66 |
|  |  | [0.20] |
| Present bias | 852 | 0.23 |
| Risk aversion | 852 | 4.19 |
|  |  | [2.05] |
| *Panel B: Sexual behaviour* |  |  |
| **HSV-2 status at baseline** |  |  |
| Negative | 731 | 59.78% |
| Positive | 731 | 29.82% |
| Equivocal | 731 | 10.40% |
| **HIV status at baseline** |  |  |
| Positive | 936 | 3.00% |
| **Perceived chance of being infected with HIV in the next 12 months** |  |  |
| High | 832 | 0.96% |
| Moderate | 832 | 3.00% |
| Low | 832 | 96.03% |
| **Age at first sex (years)** | 756 | 17.92 |
|  |  | [1.98] |
| **Currently taking PrEP** | 967 | 0.00% |
| **Correctly identified 25-29 yr old man as higher risk of HIV compared to a man 15-19 years old** | 925 | 36.50% |
| **Correctly identified 25-29 yr old man as higher risk of HIV compared to a woman 15-19 years old** | 919 | 19.70% |
| **Condom use at last sex** | 756 | 14.42% |

**Table S 2:** Baseline characteristics associated with participation in the intervention. Notes: Dependent variable is a binary indicator equal to 1 if a treatment arm individual participated in the intervention, and equal to 0 for individuals randomised to the control arm or non-compliers in the treatment arm. Estimates obtained through OLS regressions. Robust standard errors clustered at the randomisation cluster level in parentheses. *** p<0.01, ** p<0.05, * p<0.1

|  | (1) | (2) |
| --- | --- | --- |
| Age in years | 0.002 | 0.002 |
|  | (0.010) | (0.012) |
| Marital status: Never married (base) |  |  |
| Currently married | -0.082 | -0.085 |
|  | (0.110) | (0.124) |
| Divorced/separated | -0.143 | -0.115 |
|  | (0.101) | (0.120) |
| Widowed | 0.095 | 0.066 |
|  | (0.415) | (0.414) |
| Education: none/primary (base) |  |  |
| Secondary/higher | 0.115* | 0.072 |
|  | (0.057) | (0.049) |
| Economic sector: Estates (base) |  |  |
| Retail or professional services |  | 0.042 |
|  |  | (0.238) |
| Informal sector |  | 0.074 |
|  |  | (0.210) |
| Student |  | 0.040 |
|  |  | (0.193) |
| Unemployed: excluding agriculture |  | 0.110 |
|  |  | (0.189) |
| Other |  | 0.191 |
|  |  | (0.163) |
| Mean implied discount factor |  | 0.190 |
|  |  | (0.145) |
| Risk aversion |  | 0.002 |
|  |  | (0.011) |
| Perceived chance of being infected with HIV in the next 12 months: Moderate to High (base) |  |  |
| Low |  | 0.072 |
|  |  | (0.099) |
| N | 749 | 661 |

**Table S 3:** Attrition in the follow-up sample. Notes: Dependent variables are dummies equal to 1 if the information is missing for the respondent. Estimates obtained through OLS regressions Robust standard errors clustered at the randomisation cluster level in parentheses. *** p<0.01, ** p<0.05, * p<0.1

|  | (1) | (2) | (3) | (4) | (6) |
| --- | --- | --- | --- | --- | --- |
|  |  | **Outcome missing at 6-month follow-up survey** |  |  |  |
|  | **No follow-up data** | **PrEP** | **Correctly identified 25-29 yr old man as higher risk of HIV compared to a man 15-19 years old** | **Correctly identified 25-29-year-old man as higher risk of HIV compared to a woman 15-19 years old** | **Condom use at last sex** |
| Treatment group | -0.010 | -0.008 | -0.010 | -0.010 | -0.005 |
|  | (0.027) | (0.032) | (0.027) | (0.027) | (0.032) |
| N | 737 | 623 | 737 | 737 | 624 |

**Table S 4:**
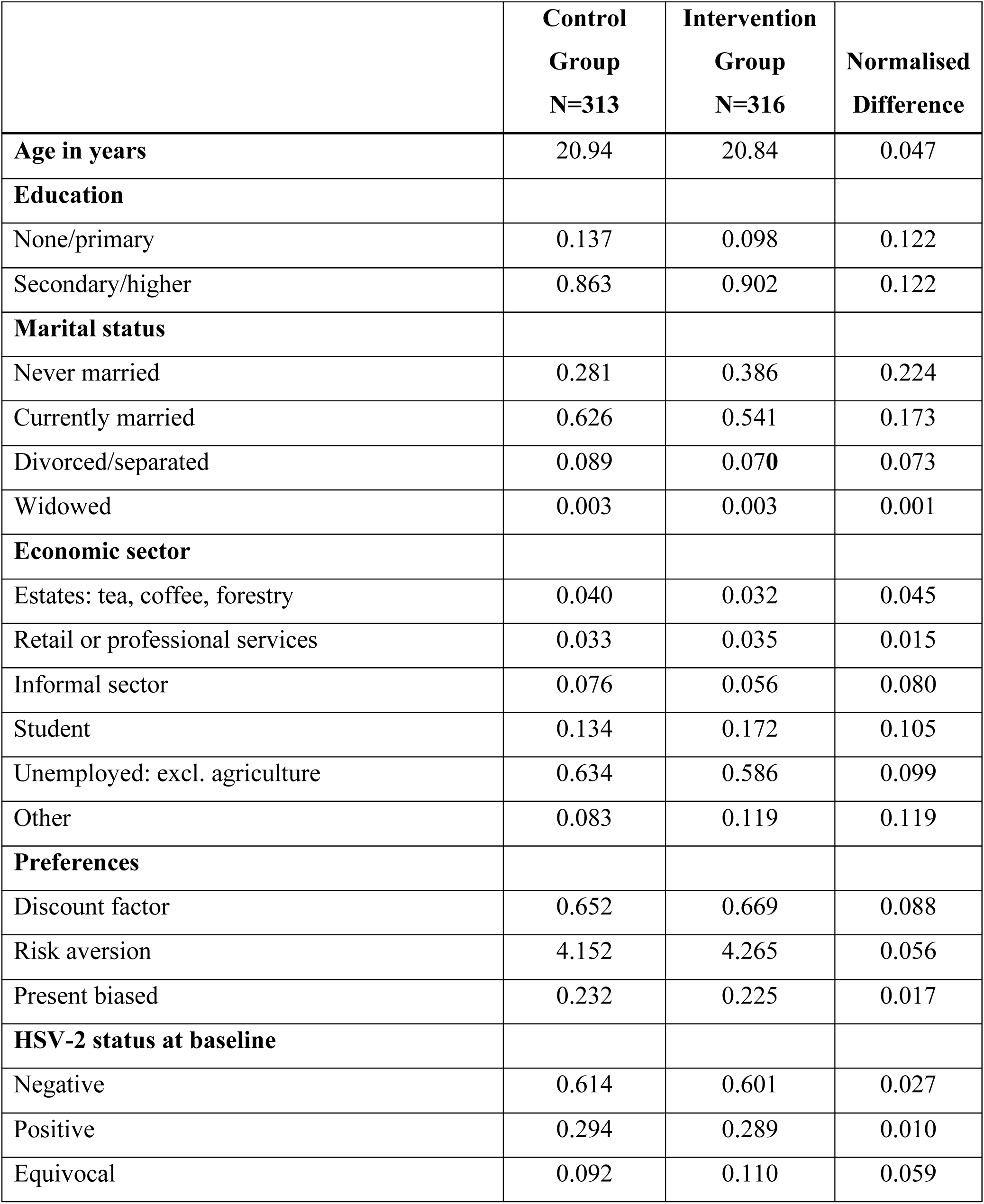
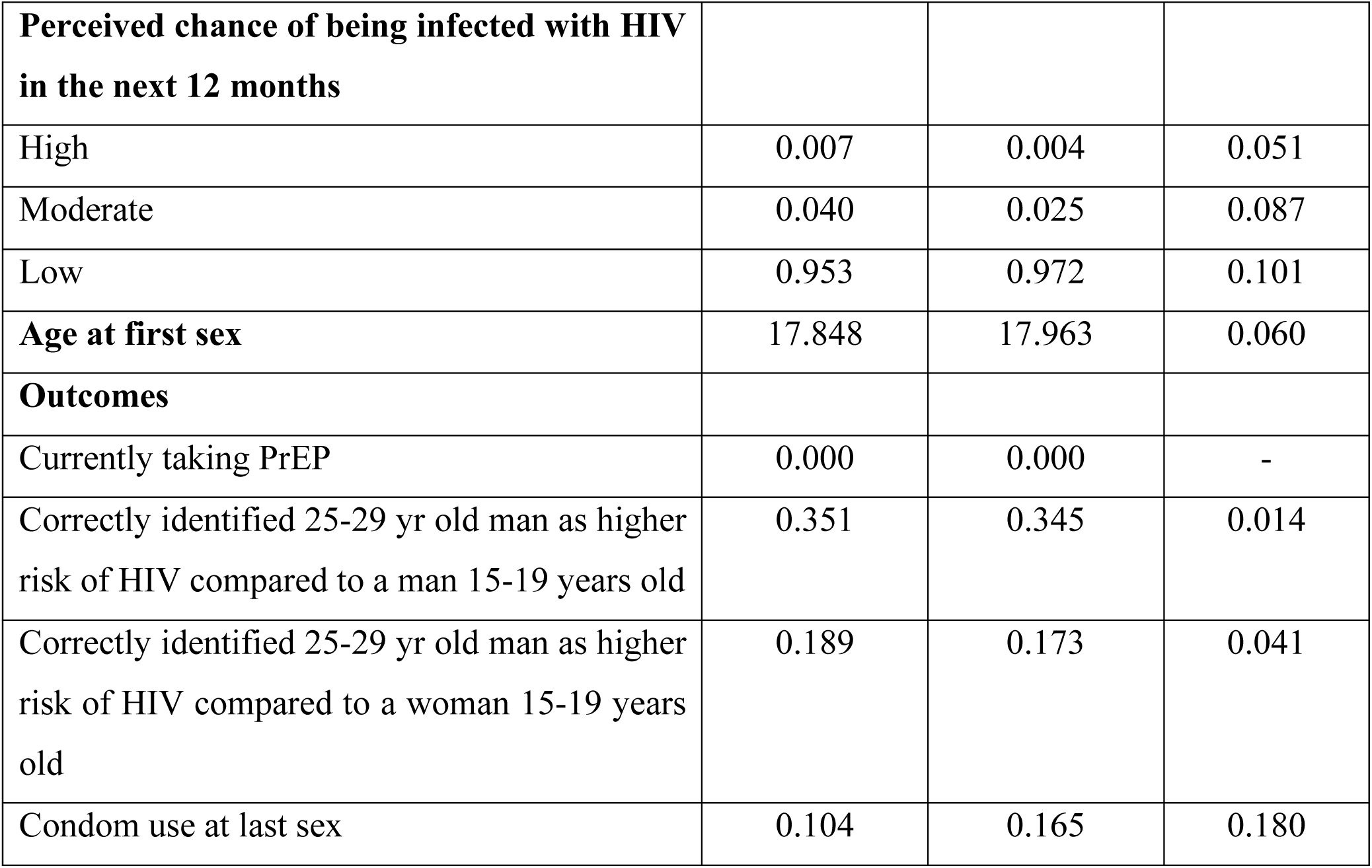
Balance checks on baseline characteristics and outcomes in the follow-up sample.

|  | **Control**<br>**Group**<br>**N=313** | **Intervention**<br>**Group**<br>**N=316** | **Normalised**<br>**Difference** |
| --- | --- | --- | --- |
| **Age in years** | 20.94 | 20.84 | 0.047 |
| **Education** |  |  |  |
| None/primary | 0.137 | 0.098 | 0.122 |
| Secondary/higher | 0.863 | 0.902 | 0.122 |
| **Marital status** |  |  |  |
| Never married | 0.281 | 0.386 | 0.224 |
| Currently married | 0.626 | 0.541 | 0.173 |
| Divorced/separated | 0.089 | 0.070 | 0.073 |
| Widowed | 0.003 | 0.003 | 0.001 |
| **Economic sector** |  |  |  |
| Estates: tea, coffee, forestry | 0.040 | 0.032 | 0.045 |
| Retail or professional services | 0.033 | 0.035 | 0.015 |
| Informal sector | 0.076 | 0.056 | 0.080 |
| Student | 0.134 | 0.172 | 0.105 |
| Unemployed: excl. agriculture | 0.634 | 0.586 | 0.099 |
| Other | 0.083 | 0.119 | 0.119 |
| **Preferences** |  |  |  |
| Discount factor | 0.652 | 0.669 | 0.088 |
| Risk aversion | 4.152 | 4.265 | 0.056 |
| Present biased | 0.232 | 0.225 | 0.017 |
| **HSV-2 status at baseline** |  |  |  |
| Negative | 0.614 | 0.601 | 0.027 |
| Positive | 0.294 | 0.289 | 0.010 |
| Equivocal | 0.092 | 0.110 | 0.059 |
| **Perceived chance of being infected with HIV in the next 12 months** |  |  |  |
| High | 0.007 | 0.004 | 0.051 |
| Moderate | 0.040 | 0.025 | 0.087 |
| Low | 0.953 | 0.972 | 0.101 |
| **Age at first sex** | 17.848 | 17.963 | 0.060 |
| **Outcomes** |  |  |  |
| Currently taking PrEP | 0.000 | 0.000 | - |
| Correctly identified 25-29 yr old man as higher risk of HIV compared to a man 15-19 years old | 0.351 | 0.345 | 0.014 |
| Correctly identified 25-29 yr old man as higher risk of HIV compared to a woman 15-19 years old | 0.189 | 0.173 | 0.041 |
| Condom use at last sex | 0.104 | 0.165 | 0.180 |

**Table S 5:** Unadjusted intention-to-treat results from cluster-level analysis. Notes: Results are unadjusted cluster-level summaries (Hayes & Moulton, 2017). Standard errors in parentheses. *** p<0.01, ** p<0.05, * p<0.1

|  | (1) | (2) | (3) | (4) |
| --- | --- | --- | --- | --- |
|  | **PrEP** | **Correctly identified 25-29 yr old man as higher risk of HIV compared to a man 15-19 years old** | **Correctly identified 25-29 yr old man as higher risk of HIV compared to a woman 15-19 years old** | **Condom use at last sex** |
| Treatment effect (ITT) | 0.041 | 0.104 ** | 0.101** | -0.010 |
|  | (0.036) | (0.033) | (0.042) | (0.087) |

**Table S 6:** Robustness checks of local average treatment effects. Notes: All models estimated using 2SLS including community fixed effects. ^@^Covariates included are age, education, marital status, perceived risk of HIV infection in the next 12 months, present bias, risk aversion, mean discount factor and a binary indicator of whether the individual was a consistent respondent to the time preference questions. Robust standard errors clustered at the randomisation cluster level in parentheses. *** p<0.01, ** p<0.05, * p<0.1.

|  | (1) | (2) | (3) | (5) |
| --- | --- | --- | --- | --- |
|  | PrEP | Correctly identified 25–29-year-old man as higher risk of HIV compared to a man 15-19 years old | Correctly identified 25–29-year-old man as higher risk of HIV compared to a woman 15-19 years old | Condom use at last sex |
| Baseline outcome and covariates | 0.058** | 0.139*** | 0.130*** | -0.047* |
|  | (0.027) | (0.028) | (0.030) | (0.025) |
| N | 517 | 629 | 598 | 487 |
| Baseline outcome and all covariates<sup>@</sup> | 0.061** | 0.134*** | 0.144*** | -0.055** |
|  | (0.028) | (0.030) | (0.034) | (0.024) |
| N | 457 | 558 | 530 | 427 |

**Table S 7:** Robustness checks of local average treatment effects without clustering and wild restricted bootstrap standard errors. Notes: All models estimated using 2SLS including community fixed effects and covariates age, education and marital status. Robust standard errors without clustering in parentheses with accompanying conventional p-values: *** p<0.01, ** p<0.05, * p<0.1. p-values from wild restricted efficient bootstrap estimated from 1000 replications are presented in brackets.

|  | (1) | (2) | (3) | (5) |
| --- | --- | --- | --- | --- |
|  | **PrEP** | **Correctly identified 25–29-year-old man as higher risk of HIV compared to a man 15-19 years old** | **Correctly identified 25–29-year-old man as higher risk of HIV compared to a woman 15-19 years old** | **Condom use at last sex** |
| Treatment effect | 0.058*** | 0.136*** | 0.135*** | -0.025 |
| Robust standard errors without clustering | (0.022) | (0.052) | (0.047) | (0.038) |
| p-values from wild restricted efficient bootstrap standard errors | [0.003] | [0.012] | [0.007] | [0.518] |
| N | 517 | 583 | 583 | 516 |

## Footnotes

1 The experiment was pre-registered on clinicaltrials.gov (NCT03565575). The statistical analysis plan was registered with the funder appointed Data, Safety and Monitoring Board during baseline data collection. The analysis plan was extended to include local average treatment effects during routine monitoring of enrolment numbers

2 In the appendices we present unadjusted intention-to-treat (ITT) estimates – including all women in the communities meeting the study inclusion criteria, regardless of whether they were invited to the intervention. Since this is an individual level intervention, we expect these community wide ITT results, to be weaker and less precise than the treatment effect on the participants. For ITT estimates, we use a two-stage approach based on analysis of cluster-level summary measures of the outcomes. This approach is standard for the analysis of cluster randomized trials with less than 15 clusters per treatment arm (Hayes and Moulton, 2017).

3 The analysis by HSV-2 status was not pre-specified in the study analysis plan.

## Notes

### Competing Interest Statement

The authors have declared no competing interest.

### Clinical Trial

NCT03565575

